# Stakeholders’ perceptions of DNDi’s interventions and collaboration in the Democratic Republic of Congo from 2005-2023: a qualitative study

**DOI:** 10.64898/2026.02.12.26346130

**Authors:** Thérèse Mambu, Eric Mafuta, Benito Kazenza, Gabriela Costa Chaves, Mona Regad, MBO Florent, Eric Stobbaerts, Chirac Bulanga

## Abstract

This study examines the impact of the Drugs for Neglected Diseases initiative (DNDi) on health research capacity and health system strengthening in the Democratic Republic of Congo (DRC) from 2005 to 2023. Using a qualitative approach with semi-structured interviews, stakeholders and beneficiaries shared their perceptions of DNDi’s interventions. The analysis, grounded in an integrative model of organizational performance, found that DNDi’s efforts significantly enhanced clinical and operational research capacity, improved healthcare infrastructure, and fostered knowledge exchange among health practitioners. Notably, the partnership contributed to reduced morbidity and mortality from sleeping sickness through the development of safer, more effective treatments such as nifurtimox-eflornithine combination therapy (NECT), fexinidazole, and acoziborole. DNDi’s support also enabled healthcare providers to expand research capacity beyond sleeping sickness, promoting collaboration and knowledge transfer between institutions. Overall, stakeholders reported positive outcomes for patients, communities, and practitioners, highlighting DNDi’s role in building sustainable research networks and enabling environments for innovation in resource-limited settings. The study underscores the importance of continued investment in research capacity and collaborative partnerships to address neglected diseases and strengthen health systems in low-resource contexts.

## 1. Introduction

The Drugs for Neglected Diseases initiative (DNDi) was created in 2003 by MSF and research institutions from several countries in the North and South (Brazil, France, India, Kenya, Malaysia) in response to the frustration of clinicians and the despair of patients faced with drugs that were ineffective, dangerous, unavailable, unaffordable, or that had simply never been developed. Only 1.1% of the 1,393 new medicines approved between 1975 and 1999 were intended for neglected diseases (1)and this fatal imbalance persisted: between 2000 and 2010, only 4% of the 850 new medicines and vaccines developed were for neglected diseases, even though they affect 1 in 5 people globally today (2,3)].

Human African trypanosomiasis (HAT), also known as ‘sleeping sickness’, is one of the first neglected diseases that DNDi focused on. This parasitic disease is caused by a trypanosome transmitted by the tsetse fly, which, depending on the species of the parasite, can take two different forms: Trypanosoma brucei gambiense and Trypanosoma brucei rhodesiense. The disease progresses in two phases: haemolymphatic and meningoencephalic. If left untreated, the parasite crosses the blood-brain barrier and invades the central nervous system, causing an advanced stage of sleeping sickness. During this phase, people develop neuropsychiatric symptoms such as sleep disorders, confusion, lethargy, and convulsions. Without access to a therapeutic option, sleeping sickness is usually fatal. HAT occurs in several African countries, with differences in incidence between countries and regions. It is estimated that between 2018 and 2022, the Democratic Republic of Congo (DRC) accounted for 61% of reported cases, with an average of 522 cases per year. Thanks to control efforts, the annual number of cases reported decreased to fewer than 2,000 in 2017 and 1,000 by 2018, However, the situation was very different in 1998, when 40,000 cases were reported and prevalence in some villages was as high as 50% in countries such as Angola, DRC, and South Sudan (4). According to the WHO, HAT caused the loss of around 1.5 million disability-adjusted life years (DALYs) in 2002. In DRC, the cost to each household was equivalent to 5 months’ income. The total number of DALYs related to HAT was 2,145 and HAT control interventions prevented 1,408 DALYs. The cost per DALY averted was 17 USD (5). The cost of HAT control is considered to be low, while the economic benefits for those affected by the disease are high, justifying the investment.

DNDi’s mission is threefold: innovation to save lives through research and development of treatments needed for neglected patients, the development of inclusive and sustainable solutions by working closely with partners in low- and middle-income countries (LMICs), and advocacy by campaigning for policy changes in favour of a fairer and more equitable research & development (R&D) system. DNDi is committed to working with the expertise present in endemic countries to strengthen existing research capacity.

Since 2003, DNDi has supported regional clinical research platforms to identify and assess patient needs and gaps in R&D, strengthen and sustain clinical research capacity, facilitate access to new treatments, and advocate for a policy and regulatory environment favourable to needs-driven R&D (6).

DNDi became involved in HAT because the treatments available at the time (melarsoprol or arsobal) were toxic and caused serious adverse events such as arsenical encephalopathy. The organisation established an office to enable the development of clinical trials in 2005 in the DRC, since it was the country most affected.

DNDi then took on the mission of developing effective, high quality, safe medicines for patients, by developing infrastructures and clinical research capacities in partnership with the local scientific and medical communities. In 2005, DNDi, together with other partners and programmes involved in the effort against HAT, created a regional platform for this disease, the Human African Trypanosomiasis Platform (or HAT Platform), the overall aim of which was to strengthen clinical and operational research capacities and methodologies in endemic countries (7). Nearly 20 years on, the time has come to assess, from the perspective of DNDi’s partners, the contribution that the DNDi partnership has made to solving the challenges of neglected tropical diseases in the DRC, and to draw lessons from this.

This case study follows a previous analysis conducted by DNDi, which investigated how the development of clinical research capacities in an LMIC contributed to strengthening its healthcare system, focussing on DNDi’s work in DRC using a WHO model based on the six pillars of the health system (8). The study was conducted and coordinated by the Kinshasa School of Public Health and focuses on the perceptions of partners involved in DNDi projects in the DRC between 2005 and 2023.

This study is part of DNDi’s strategic objective to sustain and develop clinical research networks and drug optimization programmes in LMICs. The aim of this study is to understand partners’ perceptions of the impact of the work carried out to strengthen the national health system and to implement an environment conducive to scientific research and development in the DRC. This study aligns with the second major pillar of DNDi’s mission, which focuses on inclusive and sustainable solutions in collaboration with partners in LMICS (9). This retrospective study describes stakeholders’ perceptions of the development of research capacity and the quality of collaborations that took place in the DRC between 2005 and 2023.

### DNDi trajectory in the DRC (2005-2023)

This section aims to briefly contextualise the interventions implemented by DNDi and its partners in DRC in order to better understand partners’ perceptions of the partnership, which are presented in subsequent sections.

Since 2005, DNDi has been developing projects in collaboration with a range of partners in DRC. These projects initially focused on HAT and, more recently, have extended to onchocerciasis (river blindness), COVID-19, and HIV. The clinical development strategy for new treatments was accompanied by a public health vision aimed at controlling, and then eliminating, the disease in this region, in collaboration with numerous partners. In parallel with these research projects, DNDi was seeking to develop research capabilities in resource-constrained settings, and in co-developing know-how and knowledge in the scientific field of clinical development.

For HAT, the initial strategy involved developing a tolerable and effective combination of drugs: nifurtimox-eflornithine combination therapy (NECT), which replaced the highly toxic treatment melarsoprol. But the logistical challenges of providing care in hard-to-reach areas (where most patients are) pushed DNDi to think further about how to simplify treatment for physicians and patients, which led to the development of an oral treatment: fexinidazole. In 2012, a study was launched with this easy to manage pill, which is taken orally for 10 days to treat early-stage sleeping sickness. This drug is a milestone along to path to eliminating sleeping sickness. Yet to simplify the treatment further and enhance the well-being of patients, in 2016 DNDi developed acoziborole, a single-dose treatment for all stages of the disease. Acoziborole is a game-changing innovation in terms of efficacy and tolerance.

These projects entailed clinical trials, which are needed in order to obtain marketing authorization, most of these were carried out as part of the HAT Platform during the final stage of the drug development process. The HAT Platform was created in 2005 and is a network of 120 members from more than 20 institutions (universities, NGOs, research centres, government institutions) which aims to develop and strengthen clinical research capacities and provide a forum for sharing information and connecting endemic countries. It has conducted various activities in connection with clinical trials, including the development of specific methodologies for sleeping sickness, while also working on the administrative and regulatory barriers within the health system by building capacity at individual (various training courses) and organisational levels (infrastructures, equipment, management systems).

The main objectives of this platform were to strengthen research capacity, improve the research environment, support ethical committees and operational research, and develop partnerships and a communication plan. The aim was to provide endemic countries with new screening and diagnostic tools and effective, non-toxic, preferably oral, treatments for both stages of the disease.

Prior to the start of the clinical trials, technical support was provided to the ethical committees in the endemic countries where the clinical trials were to be carried out. This support was later extended to regulatory and pharmacovigilance bodies, to enable them to monitor and report adverse drug events using tools developed by the WHO and the Uppsala Monitoring Centre (UMC). DNDi has carried out its activities through the HAT Platform, of which it is the main sponsor.

## 2. Materials and Methods

### 2.1 Theoretical framework

To optimise the possibility of answering the research questions, an integrative model of organisational performance was used to develop the variables. The model designed by Sicotte et al. is ‘based on a very general vision of the functions that any organisation must fulfil in an environment’ (10)and is closely inspired by Parsons’ Theory of Social Action. It specifies the four essential functions that an organisation must constantly maintain in order to survive. These models illustrate an aspect of organisational performance and the functions essential to the survival of organisations. The analysis also included partners’ perceptions of the impact of DNDi’s work on aspects of health systems strengthening (HSS) (11) and research capacity strengthening (12).

### 2.2 Scope, type, and period of study

The study included 14 sites in the five provinces of DRC where DNDi is active, namely Kwilu, Mai-Ndombe, Kasaï Oriental, Kasaï Occidental, and Kasaï Central. Most of the clinical trials on HAT have been conducted in these provinces, which report more than 70% of HAT cases in the DRC.

This case study is based on semi-structured interviews with DNDi partners, providers at national and provincial coordination levels, health zones, and community leaders in the regions where DNDi interventions have been carried out.

### 2.3 Sampling

The study population included management from the Ministry of Public Health who have worked in collaboration with DNDi on projects at the national and provincial levels (in particular the Heads of the Provincial Health Division (CD) and the Medical Officers of Health (MCZ) in the supported provinces); managers from the National Programme for the Control of Human African Trypanosomiasis (PNLTHA), in particular the Provincial Coordinating Physicians (MCP) in the provinces concerned; people who have worked in the clinical research sites as investigators or lab technicians and DNDi managers in DRC and Guinea (where DNDi has been conducting clinical trials since 2017); beneficiaries (patients, providers); research partners (ITM Antwerp, scientists); the laboratory network (Institut National de recherche biomedicale - INRB); and other technical and financial partners (PATH, PDSS/World Bank). At least two participants in each category were included in the study. Respondents were selected with a view to ensuring diversity in terms of professional occupation, but the main eligibility criterion was participation as stakeholders in or as beneficiaries of DNDi interventions.

### 2.4 Data collection

The data for this study were collected mainly through semi-structured interviews with key informants and a literature review. Study participants were recruited between 20^th^ March – 10^th^ April 2023. Their consent was collected at that time by the investigators (consent recorded). Interviews were conducted by telephone between 20^th^ March – 21^st^ April 2023 and were recorded on a voice recorder. The participants’ consent was renewed at that time and recorded.

### 2.5 Variables of interest

Data were collected about stakeholders’ perceptions of the development of technological capacity and technical competencies, as well as the quality of the collaboration. This was defined as the way in which stakeholders (service providers and communities (community leaders, partners)) view DNDi’s achievements in DRC since 2005, and the development of capacity during collaborative exchanges with stakeholders. In addition, the following information was collected: the stakeholder’s perception of clinical research and of the benefit that this knowledge and associated network could provide in the future; lessons learned about the relevance of DNDi activities and about their agenda at the national and international level; information from stakeholders about the development of capacity and competencies due to exchanges with DNDi, about the establishment of a national ecosystem for innovation and access to treatment, and about the quality of care after the transformation of facilities.

### 2.6 Data processing and analysis

The data from the semi-structured interviews and the document review were analysed using a deductive approach based on Sicotte’s model and content analysis (10). Content analysis consisted of listening to the recordings several times before producing a literal transcript of the interview, respecting the verbatim whose meaning was sought during the survey. A few transcripts were then selected by respondent category to enable a coding guide to be drawn up, which was used to organise the data using Atlas-ti software. Before the synthesis, a data analysis matrix was drawn up to identify the divergences, similarities, and/or convergences between the respondents. A summary was produced and an interpretation proposed based on the data collected and the information drawn from them.

### 2.7 Ethical considerations

The study protocol was submitted to the Ethics Committee of the School of Public Health of the University of Kinshasa and received approval under number ESP/ CE/026/2023. Participation in the study was subject to the respondent’s written informed consent. The study did not involve any major risks for the participants and the data were anonymised to preserve confidentiality. No direct benefit was obtained by the participants.

## 3. Results

**Table 1** provides the sociodemographic characteristics of the 32 interviewed participants and their position in relation to the various interventions undertaken in the projects developed by DNDi.

**Table 1.**
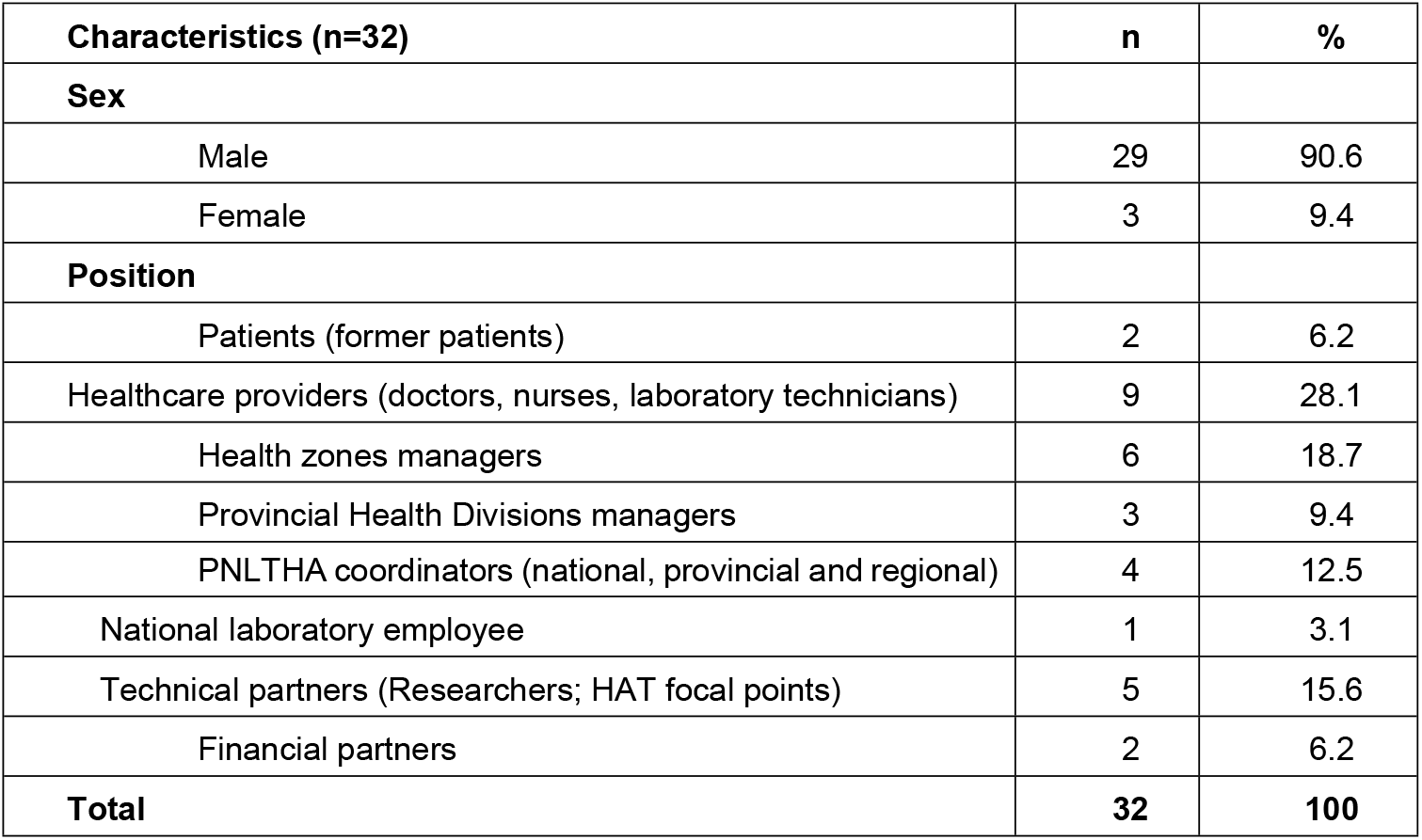
Sociodemographic characteristics of respondents.

| Characteristics (n=32) | n | % |
| --- | --- | --- |
| **Sex** |  |  |
| Male | 29 | 90.6 |
| Female | 3 | 9.4 |
| **Position** |  |  |
| Patients (former patients) | 2 | 6.2 |
| Healthcare providers (doctors, nurses, laboratory technicians) | 9 | 28.1 |
| Health zones managers | 6 | 18.7 |
| Provincial Health Divisions managers | 3 | 9.4 |
| PNLTHA coordinators (national, provincial and regional) | 4 | 12.5 |
| National laboratory employee | 1 | 3.1 |
| Technical partners (Researchers; HAT focal points) | 5 | 15.6 |
| Financial partners | 2 | 6.2 |
| **Total** | **32** | **100** |

Study respondents were drawn from two groups: beneficiaries and technical and financial partners. Beneficiaries included the provincial health manager, service providers, and former patients, while technical, financial, and research partners were those mainly involved in the screening and management of HAT cases. These included experts, laboratories, provincial and central coordinators of the PNLTHA, and the HAT focal points of the various technical and financial partners. Other technical partners, including the HAT focal points of the various technical and financial partnership (TFPs) and research structures, were also responsible for coordinating activities through the various meetings and platforms, and for monitoring and pharmacovigilance (for research structures and laboratories) of the various molecules used in clinical trials. By considering these categories, it was possible to capture different perspectives on the statements made about clinical trials implemented by DNDi in the DRC.

### 3.1 Respondents’ perceptions of interventions

#### 3.1.1 Achieving institutional goals

The respondents felt that the interventions undertaken by DNDi in different target zones had contributed considerably to improving the health systems in the target sites. These interventions have also enabled the beneficiaries, i.e. the provinces, health zones, and health facilities, to achieve their organisational goal of reducing the morbidity and mortality of HAT. The health facilities have benefited from this basic strategy, especially from screening for sleeping sickness, correct treatment of HAT, and control of the tsetse fly.

#### 3.1.2 Impact on the healthcare system

According to the respondents, the facilities in the various intervention zones have benefited considerably, in the form of rehabilitation of certain health facilities, supply and use of laboratory equipment, improvement of the energy supply, and provision of various inputs to the health facilities to improve patient care (such as medicines and nutritional support). In addition, some respondents, particularly health care providers, mentioned financial motivation in the form of a monthly performance bonus (a financial incentive) as being a big help in achieving the organisation’s (or structure’s) objectives. Some respondents said that the various training courses (knowledge acquisition and qualifications) organised for providers by DNDi had enabled them to be effective in providing quality care to patients. Respondents added the improvement in working conditions for mobile units (operating procedures).

> *“It’s a partner who has come to support us, who has added something to what we were doing. And the management of HAT, you know, first of all the health zone (Name of the health zone) is one of the districts in the country, even in Africa, that provides the highest number of HAT cases. So with our own resources, coming from our government and the community, we couldn’t do what DNDi has done, such as rehabilitating the laboratory and even the case-management centre. We already have a good laboratory with equipment that enables us to detect trypanosomes, which has enabled us to achieve our diagnostic results and to treat our patients”. (BF_08; PZS)*.

#### 3.1.3 Influence on care

Respondents unanimously affirmed that the research project and the interventions implemented by DNDi in the fight against HAT had a big systemic impact on the health of the population, community perceptions, health services, and health personnel. In terms of the health status of the general population (or target populations), respondents stated that the interventions linked to clinical trials enabled a considerable improvement in the final outcome of patients suffering from HAT by reducing mortality. The interventions have also increased the number of healthy individuals of working age in the community and improved other indirect benefits, going beyond patient care by improving hospital waste management and the healthcare environment.

Secondly, DNDi’s interventions have helped to facilitate diagnosis, to develop a safe oral treatment that does not require lumbar puncture, and to facilitate the operation of mobile units travelling from village to village for mass screening. These medical interventions, carried out as part of the clinical trial, have led to an increase in the medical knowledge of those involved in the health system.

Respondents also reported that DNDi has facilitated community participation by testing the population and organising activities to accompany patients to treatment centres. These initiatives have created a relay system within the community, transforming its members into agents of change and disease control.

Thirdly, according to the respondents, DNDi interventions helped to minimise the financial risk associated with access to care for patients. From the perspective of the beneficiary population, the provision of inputs for the treatment of HAT, the management of co-morbidities, and nutritional support for patients were the facilitators to overcoming the financial barrier to access to care, which significantly reduced the cost of services provided to patients. The fact that these interventions were given free of charge (according to the ethical framework for any clinical trial, as defined by the standards of good clinical practice (GCP) and the Declaration of Helsinki) was considered by the respondents to be a key element of their success. Some respondents stated:

> *“All HAT patients who are treated don’t even pay a franc to the hospital for their medicine. What’s more, while they’re in hospital, they’re taken care of in relation to the catering. That is already a gain in relation to finance. (BF_02; PSS). “Yes, because at the moment, there are no more therapeutic failures. So DNDi has still helped us to achieve our results, because when there was failure, there were also new cases. If a person is not treated properly, the fly can still bite them and bite a healthy person, so transmission continues”. (PTF_02)*

The study explored the responsiveness of health services. Respondents said that the active screening strategy using mobile units staffed by investigators was a determining factor in the responsiveness of the health services. Respondents recognised that the patient transport system and the availability of all the inputs needed for treatment also contributed to the responsiveness of the health services. The complementarity between the clinical research capacities strengthened by DNDi and the improvements made to DRC’s healthcare system is essential to understanding the interactions and convergences that have made it possible to improve patient care.

“The mobile units would go to places where we couldn’t go, and detect the sick in the depths of the countryside, and they would be taken to a hospital. And they were treated free of charge, and after the check-ups they were declared cured… the health of the whole community is improved”. (BF_01; PZS)

#### 3.1.4 Impact on clinical research capacity

Respondents noted that the interventions initiated by DNDi have enabled providers to improve their skills in the management of neglected tropical diseases (NTDs), especially HAT, and other diseases in general. With DNDi’s support, staff have been recruited and trained to conduct clinical research in line with international standards. The majority of providers approached as part of this study mentioned DNDi’s contribution to strengthening their capacity in GCP. The continuing scientific and medical training provided by DNDi, and the management guides made available to providers, have served as a framework for strengthening providers’ ethical and scientific capacities regarding clinical and medical research on NTDs. In addition, the knowledge-sharing activities in health facilities for colleagues who had not taken part in the training provided an opportunity for them to improve their knowledge of NTDs and to keep up to date with the other trained providers.

> *“We were trained*…*After the training, we had to give a presentation to the whole hospital team and everyone in the hospital at least benefited from knowledge of good clinical practice and we are starting to experience this in the field with our patients”. (BF_02; PSS)*
>
> *“The staff used in the clinical trials have been trained. Their capacities have been strengthened. The rest of us have been trained in good practice and in monitoring clinical trials. So this has enabled us to have a critical mass of people who know how to implement*…*” (Research partner_02)*

Although a passion for medical research was ignited in some of the providers involved in the various interventions, these providers have not been able to increase the research capacity of their respective structures for the conduct of future clinical trials. Despite the structures’ participation in the implementation of interventions, studies and sub-studies, they have remained dependent on DNDi for their capacity to conduct these types of study, mainly due to a lack of long-term material, financial, and structural resources. In concrete terms, as the sponsor, DNDi provided the protocol, logistics, and all the funding during the studies, as well as the technical capacity that the local structures did not have, while the service providers benefited from capacity strengthening and the structures obtained specific equipment. The structures lack the capacity to conceptualise these types of studies, the connection to international research networks, and the logistics that accompany them in terms of inputs, equipment, and infrastructure, given the state of the country’s health facilities.

Nevertheless, the experience gained with DNDi is now an asset that can be used for the professional development of those involved in running the trials, and for subsequent studies within the country or internationally. For some investigators, participation in these clinical trials was also an opportunity to author scientific articles in collaboration with DNDi and the PNLTHA, and to progress scientifically (13).

> *“We think we’re a little advanced because us, for example as investigators, the training we received with the clinical trial was an international training. Today, we are capable of managing a study anywhere else. So we are able to today conduct a study even outside the country”. (BF_02; PSS)*

### 3.2 Perceptions of related spin-offs, contributors and challenges

#### 3.2.1 Quality of the partnership

Respondents unanimously reported an excellent level of satisfaction with the partnership, both for the patient population and for the providers themselves. The new treatments have several advantages, in particular a significant reduction in the side-effects associated with the previous products, which was at the root of the fear and non-use of HAT treatment facilities by patients with HAT. According to respondents, the new management strategy has increased patient attendance and adherence to treatment. One respondent said:

> *“*… *people are very happy with this initiative because you know with injectable products there are other people who are afraid to take them. But as these are tablets, they swallow them easily; previously there were people who refused to be admitted in hospitals, but with fexinidazole, which we give on an outpatient basis, you follow them at home”. (BF_04; PSS)*

Respondents also reported that the partnership with DNDi had enabled their respective organisations to carry out their missions fully during the intervention period. In concrete terms, thanks to this partnership, the organisations were able to benefit from the necessary resources, such as the construction and rehabilitation of basic infrastructure, provision of essential inputs and medicines, integrated training on NTDs for service providers, the provision of care guidelines, free-of-charge services, the sensitisation and integration of community leaders in the care process, and sufficient financial motivation. These interventions have enabled their organisations to acquire new skills, to experience an influx of people seeking services, to improve their response to the needs of the target populations, to limit brain drain, and to adapt to change, thanks to an improved working environment. One respondent said:

> *“We’ve kept in close touch with patients, we can call them, whereas before it wasn’t possible, communication has improved, there’s the internet*… *And also, HAT patients, who were used to being treated by the mobile unit team, who didn’t come to the hospital, have started to come to the hospital. But also for those who were ashamed of HAT, because it was said that the patient was either bewitched or a sorcerer, and people were hiding*… *because the disease was no longer clandestine and people knew that they could be treated and cured. We used to use binocular microscopes, but with DNDi’s microscopes, which have cameras, everyone involved in the study can see the trypanosome - it’s a huge change. (BF_01; PZS)*

#### 3.2.2 Productivity of structures

The respondents affirmed that the previously mentioned actions in support of clinical trials, of which the structures have been beneficiaries, have increased their productivity in terms of the volume of services offered and that, above all, they have improved the quality of care and services offered. In addition, the various research projects have identified effective and innovative therapeutic regimens that significantly reduce the adverse effects in patients, which has helped to guide new policies for the management of NTDs, particularly HAT, and to contribute to a change of scenario for health research in the DRC. One participant said:

> *“Because back home, HAT was a disease thought to be linked to witchcraft*… *And people had to be treated with NECT, which required a lot of injections and a lot of techniques*… *Few people were attracted to treating these patients*… *We switched from injectable to oral medicines. But also with regard to screening, the means of screening were limited, but with the clinical studies, even in remote areas, there were the necessary means*,, *to go and find patients even in the depths of the countryside, screen them, take them to hospital and ensure that they received a single-dose treatment”. (BF_02; PSS)*

This description of the therapeutic progress made for HAT demonstrates the impact of improving clinical research capabilities and strengthening the healthcare system, while keeping patients’ needs at the heart of the intervention.

#### 3.2.3 Contributors to success

According to the beneficiaries, the working conditions created by DNDi, characterised by transparency, willingness of project coordinators and supervisors to listen, a friendly atmosphere, the availability of communication tools, such as the internet, and the absence of input shortages, contributed to this level of satisfaction. Other contributors reported by the respondents include the financial motivation for service providers and the practical working environment (water, electricity, technical support, medicines).

> *“We are satisfied with DNDi’s sincerity. Everything they taught us at the beginning, we saw as such. But also the simplicity with which we worked, we didn’t feel a disconnect between the teachers and us. We really felt that we were a team and we really valued our work through the partnership with DNDi”. (BF_01; PZS)*

#### 3.2.4 Interaction and interconnection

Within the framework of the activities implemented by DNDi, effective coordination required the participation of various players at different levels, from the health facility to the coordinating investigator, via the provincial coordination teams. In practical terms, the organisations benefited from training, supervision, and monitoring activities during the intervention periods. In fact, this collaboration enabled the organisations not only to maintain their values and standards, but also to develop other values aimed at improving the provision of services to the community. These include values such as good reception of patients, treatment based on guidelines (standard operating procedures, SOPs), improvements to the care environment (waste treatment, rehabilitation of wards, provision of equipment) and dedication to patients, which is often hampered by the lack of adequate remuneration for care staff.

> *“There are many objectives and activities at the hospital, not just the treatment of HAT. The aim of any health structure is to provide patients, whatever their pathology, with good conditions, and to treat them correctly for their problem. The refurbishment of the wards, and the equipment provided by DNDi, means that patients with other pathologies can also be received and cared for. The lighting, for example, is not just for HAT patients, but for the whole hospital*… *so indirectly, it also solves the problems of other pathologies. In the laboratory, we can see the parasite, we’re sure of the diagnosis, and the treatment that follows is a very good one. As a hospital, our aim was to support the research, and the resources that DNDi provided enabled us to support the study right through to the end. (BF_02; PSS)*.

In addition, this collaboration has encouraged the transmission of knowledge and the sharing of experience between the various stakeholders. This was facilitated by a strategy of weekly meetings.

> *“Exchanges are maintained through meetings, whether online or face-to-face, and some people have attended conferences*… *National and international exchanges are intense, not only with DNDi but also with other organisations. And those who have taken part in clinical trials keep close links and when there is an opportunity for some kind of training, people share it”. (BF_01; PZS)*
>
> *“Yes, there are exchanges of knowledge between international and national levels, yes, because particularly for investigators, they have the opportunity to attend international conferences, and they have investigators’ meetings, where all levels get together to evaluate the activities carried out, so there’s that, and it allows contact and helps providers to relax. (PLTHA_05)*

Respondents also mentioned the key role played by the HAT Platform in achieving the expected results. It was responsible for training and conferences, guidance on diagnosis and ensuring a paradigm shift on NTDs in the community through its discussions with political and administrative authorities. It was also responsible for ensuring the effective implementation and smooth running of the clinical trials.

> *“The platform organised most of the training courses and conferences* … *it played a role in capacity building, but also in leadership, by explaining to the political and administrative authorities the benefits of keeping the people we trained in their places of work to carry out the studies”. (BF_01; PZS)*

#### 3.2.5 Challenges associated with carrying out DNDi interventions

Some respondents reported that this collaboration led to frustration among providers in the implementing health facilities, due to the difference in remuneration between care providers involved in clinical trials and other staff who were not involved. While this should be taken into account when considering how to further improve the working environment, it is a situation that is always observed when partners provide support to national health facilities, because the limited resources cannot stretch to cover all the staff:

> *“There is cohesion among the staff in the study because we were trained together and we now share the same values. But with all the members of the organisation, there have been frustrations*… *In our environments, people earn too little money and sometimes those who are not in the study can campaign for a team member to be transferred so that he or she can be replaced*…*”. (BF_01; PZS)*

Another major challenge faced by those involved was the change in the community’s perception of the disease. It was initially difficult for providers to convince patients of the benefits of new products and to obtain their consent to participate in clinical trials, but a strategy of involving and training community leaders and community health workers has helped the community to embrace the project.

> *“Yes, talking about challenges, it all depends on resistance, of people who still believe it is a bad spell, the churches also pose problems because they give out messages that this illness is witchcraft, that people should pray instead of going to hospital, and that there are customs too. But I think that as time goes by, people understand that it’s just an illness”. (BF_03; PSS)*

Nevertheless, respondents expressed concern about the sustainability of DNDi’s achievements in strengthening the pillars of the healthcare system. One participant said:

> *“We ask God to continue such initiatives, not to leave us, because if DNDi leaves us today, we will be abandoned because it has removed poverty. Me talking to you now, I’ve been working for 33 years, I’m not a salaried employee, but I’ve sent my children to school through DNDi*…*all our compatriots and service providers together congratulate these initiatives for taking care of us in terms of financial resources”. (BF_04; PSS)*

In addition, the project has not led to widespread protection of the population’s health. The focus was essentially on patients suffering from HAT, and only these patients had access to the various inputs. On the other hand, rehabilitation of basic infrastructure and improvement of the health environment can be considered as achievements that provide indirect benefits for other types of patients.

> *“The general feeling no, it’s as I told you, they are more focused on their clinical trial activities and do not allow for a generalised action. I’ll give you an example of how now, at facility level, only those people who have previously been included in their studies have access to their equipment, You can already see that this is a limiting factor, which does not allow general improvement but which will allow improvement for those who are included in the study, possibly for cases where a patient suffering from malaria comes to the ward and sleeps on that bed, so it’s a bit indirect*.
>
> *“* …*the trypanosomiasis project is a project that is absolutely not a priority for the Ministry of Health in the DRC, and rightly so, because despite our efforts to test more than two million people a year, in which DNDi has played a role, in the end very few people are affected and there are many other health problems that are much more important for a country like the DRC, and so it is logical that the resources for health, from the Ministry of Health, should be directed towards problems other than trypanosomiasis”. (PTF_04)*

## 4. Discussion

### 4.1 Objectives and results of the partnership with DNDi

#### 4.1.1 DNDi objectives

The study explored the perceptions of DNDi’s partners of the implementation of clinical trials in DRC, including patients, health professionals, and health managers at sub-national and national levels, reflecting a diverse range of positions across the health system, and giving them the opportunity to provide different perspectives on the effects of the partnership. The partnership has enabled screening and treatment activities, as well as ongoing research into better treatment options over time.

#### 4.1.2 Results obtained

The objectives of the interventions were perceived as being achieved by the reduction in HAT morbidity and mortality over the years. Since 2000 there has been a decrease in the number of HAT cases in endemic countries: from over 25,000 in 2000 to over 15,000 in 2005. During the NECT development period, from 2005 to 2009, the number of cases fell again significantly, reaching less than 10,000 in 2009. The sustained reduction in cases, together with the availability of better treatment options, has made HAT one of the NTDs targeted by the WHO for elimination by 2030 (14)

### 4.2 Impact on patients and the community

#### 4.2.2 The role of clinical trials

In the DRC, the elimination of HAT has become a tangible possibility thanks to the availability of the new treatment modalities and to the participation of patients in DNDi clinical trials: patients participating in trials and receiving care can promote these initiatives and encourage others to visit health facilities and receive treatment. The sustained efforts of mobile teams to seek out patients in remote areas have been crucial to the success of the trials and the reduction in morbidity.

#### 4.2.3 Community perception of the interventions

There was a unanimous perception that DNDi interventions had an impact on the health of the population at an epidemiological level, illustrated by people’s sense of well-being and productive capacity. The community was involved in the active case-finding strategy and in following up positive cases to seek treatment in health facilities.

### 4.3 Effects on the healthcare system

#### 4.3.1 Perceived improvements

Respondents perceived improvements in healthcare facility infrastructure and working conditions resulting from the partnership with DNDi, as well as in financial incentives and training for healthcare professionals. One of the building blocks of the healthcare system is the ‘health workforce’, which was perceived to have been improved as a result of the partnership.

#### 4.3.1 Inequalities and challenges

However, some interviewees mentioned that DNDi’s presence had led to inequalities within health facilities, as professionals who were part of the DNDi study were perceived to benefit from it (in terms of finance and training) more than those who were not included. This comment is in line with the abundant literature about the inequalities that can arise when Western partners intervene in the healthcare systems of LMICs, calling for codes of conduct to be put in place and for sustainable and equitable partnerships with local stakeholders (15,16)

#### 4.3.3 Strengthening research capacity and developing an innovation ecosystem in the DRC

The partnership was seen to strengthen research capacity in a number of ways, including by improving infrastructure and developing the skills of professionals working on NTDs, by providing training in research and continuing education, and by co-publishing in scientific journals. To maintain and retain professionals with the skills to implement clinical trials, the future of a research program must be considered and incentives must be established to allow implementation of further clinical trials for other health needs in DRC. Strengthening research capacities, combined with the construction of the necessary physical infrastructure and the development of relationships with stakeholders in the healthcare system, has resulted in a favourable ecosystem for clinical trials. In May 2021, DNDi launched its first onchocerciasis trials in the DRC, using the Kimpese and Masi-Manimba sites for two studies. The existing infrastructure, as well as the knowledge and expertise of investigators and staff previously trained and involved in HAT studies, were essential to enabling these new trials. This is a good example of how building research capacity contributes directly to the development of an innovation ecosystem.

### 4.4 Partnership implementation and coordination of stakeholder involvement

The implementation of the partnership was seen to involve several players at different levels and contributed to the exchange of knowledge both nationally and internationally.

#### 4.4.1 Knowledge exchange

The HAT Platform was seen as not only contributing to the management of the disease, but also as playing a key role in promoting training and conferences, and in raising the profile of NTDs at the highest levels of government. Thanks to DNDi, the interventions undertaken have enabled the various stakeholders to achieve their goals. The development of a circuit of care, formative supervision, investigations through mobile units, regular funding of activities, meetings of different platforms, and community participation were reported as key determining factors in achieving this objective.

The interventions had a significant impact on efforts to address HAT at all levels. In terms of population health, they provided the target populations with access to effective care and recovery of their physical health. From a financial point of view, the project significantly reduced the risk for patients by covering all costs associated with treatment (hospitalisation, laboratory tests, catering, and medicines). In addition, these interventions have improved the responsiveness of health services to people in need by building the capacity of service providers, rehabilitating hospital infrastructure, developing an adequate technical platform, and ensuring the availability of inputs. Along with community engagement, these improvements have considerably strengthened HAT patients’ confidence in and support of the project’s various interventions.

The interventions implemented by DNDi have created strong links between different stakeholders. Respondents mentioned the excellent coordination of activities at all levels (health facilities, investigators, community, provincial coordination, and TFPs). The training, supervision, and monitoring activities during the interventions, as well as the coordination meetings, were decisive in strengthening collaboration between the players. This effective collaboration enabled the various stakeholders to maintain their values and standards, while also improving other services offered to the community. These included developing guidelines, improving the working environment for providers, achieving objectives, and improving the care environment for patients. In addition, this programme has improved the flow of communication and expertise between actors at the local level, represented by the PNLTHA, and a network of laboratories, researchers, and actors at the international level.

#### 4.4.2 Challenges and prospects

However, the interventions also raised some problematic issues: the frustration of certain providers in the implementing health facilities and the inequalities in remuneration for care providers involved in the trials compared to those who were not. This issue needs to be resolved in order to further improve the working environment. A plan for sustaining what has been achieved thus far and for ensuring ownership and continuity of quality care has not been sufficiently developed and discussed with stakeholders and beneficiaries.

## 5. Conclusions

The study showed that major progress has been made through the interventions implemented by DNDi in the DRC, targeting NTDs in general and HAT in particular. According to stakeholders and beneficiaries, these various interventions and actions have made a significant contribution to improving and strengthening the various pillars of the healthcare system in the DRC (leadership, infrastructure, information, financing, staff training). The innovative aspects of patient care, with the development of mobile teams and the involvement of community leaders, were mentioned by respondents as determining factors in the success of the interventions. These actions and interventions have also fostered links between institutions (programmes, health zones, health facilities) and with international bodies (TFPs), facilitating the transfer of knowledge between different players. Despite the major contributions resulting from the interventions implemented by DNDi, various stakeholders noted that the major remaining challenge is how to sustain these achievements to ensure their long-term impact on the healthcare system.

Lessons learned from partner perspectives suggest that at least three levels should be addressed to sustain this work: for patients, interventions should be free at the point of care; for health services and professionals, continued activities need to be funded; and for health research, incentives to conduct new clinical trials in the DRC must be put in place in order to continue strengthening existing research capacity. The ongoing studies on COVID-19 and onchocerciasis illustrate how the legacy of the work on HAT can be redirected to support research into priority medical needs.

Our study illustrates that, despite these challenges, partners consider DNDi’s mission to foster inclusive and sustainable solutions to be working at country level. Over the years, DNDi has collaborated with partners at different levels in the DRC to implement clinical trials for improved treatments for HAT and create the HAT Platform, overcoming barriers within the health system whilst also improving it. The lessons learned through this work are particularly pertinent in the light of the recent call to action signed by the Heads of State and Government of the African Union (17)which includes, among other things, strengthening African public health institutions and the health workforce, as well as promoting action-oriented and respectful partnerships.

## Data Availability

The data underlying the results presented in the study are available from Prof. Thérèse Mambu, from the School of Public Health at the University of Kinshasa.

## Acknowledgement of author Contributions

Conceptualization, E.S, G.C.C, M.R, F.M, C.B and T.M.; methodology, E.S, G.C.C, M.R, F.M and T.M.; software, T.M, E.M and B.K.; validation, T.M, E.M and B.K.; formal analysis, T.M, E.M and B.K.; investigation, T.M, E.M and B.K.; resources, T.M, E.M and B.K.; data curation, T.M, E.M and B.K.; writing—original draft preparation T.M, E.M and B.K.; writing—review and editing, E.S, G.C.C, M.R. F.M and T.M.; visualization, M.R.; supervision, E.S.; project administration, F.M and C.B.; funding acquisition, E.S. All authors have read and agreed to the published version of the manuscript.”

## Informed Consent Statement

Informed consent was obtained from all subjects involved in the study.

## Abbreviations

The following abbreviations are used in this manuscript:

CD: Provincial Health Division
DALYs: Disability-Adjusted Life Years
DNDi: Drugs for Neglected Diseases initiative
DRC: Democratic Republic of Congo
GCP: Good Clinical Practice
HAT: Human African trypanosomiasis
INRB: Institut National pour la Recherche Biomedicale
LMIC: Low- and Middle-Income Countries
MCP: Provincial Coordinating Physicians
MCZ: Medical Officers of Health
NECT: Nifurtimox-Eflornithine Combination Therapy
NTD: Neglected Tropical Disease
PNLTH A: National Programme for the Control of Human African Trypanosomiasis
R&D: Research and Development
SOP: Standard Operating Procedures
TFP: Technical and Financial Partnership
UMC: Uppsala Monitoring Centre
WHO: World Health Organization

## References

1. Médecins Sans Frontières Access to Essential Medicines Campaign, Drugs for Neglected Diseases Working Group. Fatal Imbalance: The Crisis in Research and Development for Drugs for Neglected Diseases. Pecoul B, Orbinski J, Torreele E, Berman D, Moon S, Warpinski A, editors. Geneva: MSF; 2001.

2. Pedrique B, Strub-Wourgaft N, Some C, Olliaro P, Trouiller P, Ford N, et al. The drug and vaccine landscape for neglected diseases (2000-11): a systematic assessment. Lancet Glob Health [Internet]. 2013 [cited 2024 Jun 20];1(6):371–9. Available from: www.thelancet.com/lancetgh

3. World Health Organization. Neglected tropical diseases: impact of COVID-19 and WHO’s response – 2021 update. Weekly Epidemiological Record [Internet]. 2021;96(38):461–8. Available from: https://iris.who.int/handle/10665/345383

4. World Health Organization. Trypanosomiasis, human African (sleeping sickness) [Internet]. 2023 [cited 2024 Jun 20]. Available from: https://www.who.int/en/news-room/fact-sheets/detail/trypanosomiasis-human-african-(sleeping-sickness)

5. Fèvre EM, Wissmann B V., Welburn SC, Lutumba P. The burden of human African Trypanosomiasis. Vol. 2, PLoS Neglected Tropical Diseases. 2008.

6. Global networks | DNDi [Internet]. [cited 2025 Jul 24]. Available from: https://dndi.org/global-networks/

7. HAT Platform | DNDi [Internet]. [cited 2025 Jul 24]. Available from: https://dndi.org/global-networks/hat-platform/

8. Mbo F, Mutombo W, Ngolo D, Kabangu P, Mordt OV, Wourgaft NS, et al. How Clinical Research Can Contribute to Strengthening Health Systems in Low Resource Countries. Tropical Medicine and Infectious Disease 2020, Vol 5, Page 48 [Internet]. 2020 Mar 29 [cited 2024 Jun 20];5(2):48. Available from: https://www.mdpi.com/2414-6366/5/2/48/htm

9. DNDi. DNDi Strategic Plan 2021-2028 [Internet]. 2021 [cited 2025 Jul 24]. Available from: https://dndi.org/publications/2021/strategic-plan-2021-2028/

10. Sicotte C, Champagne F, Contandriopoulos AP. La performance organisationnelle dans les organisations et les soins de santé. Ruptures: Revue transdisciplinaire en santé. 1999;6(1):34–101.

11. World Health Organization. Monitoring the building blocks of health systems: a handbook of indicators and their measurement strategies [Internet]. Geneva: World Health Organization; 2010 [cited 2024 Jun 20]. xii, 92 p. Available from: https://iris.who.int/handle/10665/258734

12. Franzen SRP, Chandler C, Lang T, Samuel D, Franzen RP. Health research capacity development in low and middle income countries: reality or rhetoric? A systematic meta-narrative review of the qualitative literature. bmjopen.bmj.com [Internet]. 2017 [cited 2024 Jul 8];7:12332. Available from: https://bmjopen.bmj.com/content/7/1/e012332.abstract

13. Nkieri M, Mbo F, Kavunga P, Nganzobo P, Mafolo T, Selego C, et al. An active follow-up strategy for serological suspects of human African trypanosomiasis with negative parasitology set up by a health zone team in the Democratic Republic of Congo. Trop Med Infect Dis. 2020;5(2).

14. World Health Organization. Ending the neglect to attain the sustainable development goals: a road map for neglected tropical diseases 2021–2030 [Internet]. World Health Organization; 2020 [cited 2024 Jun 20]. 177 p. Available from: https://iris.who.int/handle/10665/338565

15. Ahmed S, Chase LE, Wagnild J, Akhter N, Sturridge S, Clarke A, et al. Community health workers and health equity in low- and middle-income countries: systematic review and recommendations for policy and practice. International Journal for Equity in Health 2022 21:1 [Internet]. 2022 Apr 11 [cited 2024 Jun 20];21(1):1–30. Available from: https://equityhealthj.biomedcentral.com/articles/10.1186/s12939-021-01615-y

16. Pfeiffer J, Johnson W, Fort M, Shakow A, Hagopian A, Gloyd S, et al. Strengthening health systems in poor countries: A code of conduct for nongovernmental organizations. Vol. 98, American Journal of Public Health. 2008.

17. Africa CDC. Call To Action: Africa’s New Public Health Order [Internet]. [cited 2025 Feb 19]. Available from: https://africacdc.org/news-item/call-to-action-africas-new-public-health-order/

